# Unmet Health System Needs in an Industrial–Peri-Urban Setting: A Qualitative Study from Namanve Industrial Park and Surrounding Communities, Uganda

**DOI:** 10.64898/2026.02.10.26345964

**Authors:** Atuhaire Justus, Kato Emmanuel, Ndeezi Grace, Prichard Denzel Kavuma, Kimwise Alone

## Abstract

**Background:** Industrial and peri-urban settings present complex health challenges driven by occupational exposures, environmental risks, and socioeconomic vulnerability. Despite ongoing health education and preventive efforts, many populations living and working in such settings continue to experience significant unmet health needs that limit wellbeing and access to care.

**Methods:** This cross-sectional qualitative study was conducted in Namanve Industrial Park and surrounding communities in Mukono District, Uganda, as part of a baseline assessment to inform a planned health education intervention. Data were collected through focus group discussions (FGDs) and key informant interviews (KIIs) involving industrial workers, supervisors, health and safety personnel, teachers, school administrators, school nurses, and community stakeholders. Data were analysed using an inductive thematic analysis approach to identify unmet health needs and related systemic gaps.

**Results:** Participants articulated multiple, interrelated unmet health needs spanning preventive and primary healthcare services, sexual and reproductive health, first aid and occupational safety, water, sanitation and hygiene (WASH), environmental health, and mental health and psychosocial support. Frequently reported gaps included limited access to routine screening and testing services, lack of essential commodities such as first aid supplies, sanitary pads, personal protective equipment, and soap, inadequate WASH infrastructure, insufficient mental health and counselling services, and structural barriers related to informal employment and poor living conditions. These unmet needs were commonly expressed through requests for materials and services, reflecting broader health system and institutional shortcomings rather than individual dependency.

**Conclusion:** The findings demonstrate that unmet health needs in Namanve Industrial Park and surrounding communities are driven by systemic and structural gaps that constrain access to basic healthcare and preventive services. Addressing these needs requires integrated interventions that combine health education with improved service delivery, essential commodities, and supportive environments. Baseline evidence from this study provides critical guidance for designing context-appropriate, sustainable health interventions in industrial and peri-urban settings.

## Introduction

Industrial and peri-urban settings represent complex health environments where occupational exposures, environmental risks, and socioeconomic vulnerabilities intersect. In such contexts, workers, students, and surrounding communities are often exposed to preventable health risks while facing significant barriers to accessing basic healthcare services. Recent evidence from low- and middle-income countries demonstrates that populations living and working in industrial–peri-urban interfaces experience disproportionate health burdens due to weak service coverage, environmental hazards, and structural barriers to care (Abejirinde, 2022).

Globally, unmet health needs remain a major contributor to preventable morbidity, particularly among populations engaged in informal or precarious employment. Studies show that workers in informal and casual labour arrangements are less likely to access preventive services, routine screening, and early treatment due to cost, time constraints, and lack of employment-linked health protection (Wang et al., 2021). In low- and middle-income countries, these challenges are further amplified by under-resourced primary healthcare systems, limited occupational health services, inadequate water, sanitation, and hygiene (WASH) infrastructure, and restricted access to sexual and reproductive health services (WHO & UNICEF, 2023).

Industrial zones located within or adjacent to peri-urban settlements often magnify these vulnerabilities, as rapid industrialisation outpaces the development of supportive health and social services. Evidence from urban and peri-urban Africa indicates that informal settlements surrounding industrial areas are characterised by overcrowding, poor sanitation, environmental pollution, and limited access to affordable healthcare, resulting in higher rates of infectious diseases, occupational injuries, and psychosocial stress (Cacciatore et al., 2025). These conditions undermine the effectiveness of health promotion efforts and contribute to delayed care-seeking and reliance on self-medication.

Namanve Industrial Park, Uganda’s largest industrial hub, and its surrounding communities exemplify this industrial–community interface. The area hosts a large population of predominantly young workers, students, and casual labourers, many of whom reside in nearby congested informal settlements characterised by poor housing, inadequate sanitation, and limited access to basic services. Although some industries and schools provide fragmented health services or occasional health education activities, access to routine preventive care, screening, occupational safety measures, and psychosocial support remains inconsistent. Studies from similar urban informal settings in Uganda demonstrate that proximity to health facilities does not guarantee access, as affordability, acceptability, and service readiness remain major constraints (Dickson-Gomez et al., 2025).

In qualitative and community-based research, unmet health needs are often articulated indirectly through requests for materials, services, or institutional support. Such requests—frequently perceived as demands for donations—can be more accurately understood as expressions of structural and systemic gaps in health service provision. Interpreting these requests as indicators of unmet need provides critical insight into where health systems fail to support basic prevention, early detection, and wellbeing, particularly among vulnerable populations such as casual workers, adolescents, and women (Ogutu et al., 2021).

Understanding unmet health needs from the perspectives of those living and working within industrial and peri-urban environments is therefore essential for designing effective, contextually appropriate interventions. Evidence increasingly shows that health education alone is insufficient when individuals lack access to essential commodities, services, and supportive environments necessary to act on health information (Abejirinde, 2022; WHO, 2023). Baseline assessments that capture unmet needs are critical for informing integrated health promotion strategies that combine education with service delivery, environmental improvements, and health system strengthening.

As part of a planned health education intervention titled “Improving the Basic Health in Uganda Using e-Contents: A Case Study of Namanve Industrial Park and Surrounding Communities,” this study was conducted to identify and explore unmet health needs affecting workers, students, and surrounding communities. Using focus group discussions and key informant interviews, the study aimed to generate context-specific evidence on gaps in basic healthcare services, preventive support, and psychosocial wellbeing. The findings are intended to inform the design and implementation of integrated, sustainable interventions that respond to both expressed needs and underlying health system challenges in industrial and peri-urban settings.

## Methods

### Study Design

This study employed a cross-sectional qualitative study design to explore unmet basic healthcare needs and broader health challenges among stakeholders in an industrial and peri-urban setting. A qualitative approach was considered appropriate because the study aimed to generate in-depth, context-specific insights grounded in participants’ lived experiences and priorities. Data were collected through focus group discussions (FGDs) and key informant interviews (KIIs) to capture both collective and individual perspectives. The study was conducted as part of a baseline assessment to inform the planned health education intervention titled “Improving the Basic Health in Uganda Using e-Contents: A Case Study of Namanve Industrial Park and Surrounding Communities.”

### Study Setting

The study was conducted in Namanve Industrial Park and surrounding communities within Mukono District, Uganda. Namanve Industrial Park is the largest industrial hub in the country and hosts a wide range of manufacturing, processing, and storage industries. The surrounding area includes densely populated peri-urban communities and several secondary schools whose students and staff interact closely with the industrial environment. This setting was selected due to its unique convergence of occupational, environmental, and social health risks, coupled with vulnerabilities related to informal settlement living conditions and limited access to affordable, quality preventive and primary healthcare services.

### Study Population and Participant Selection

Participants were purposely selected based on their lived experience, institutional roles, or direct interaction with health needs and health services within industrial, school, and community contexts. The study population comprised industrial workers and supervisors, health and safety personnel, teachers and school administrators, school nurses and staff involved in student welfare, and other community-based stakeholders involved in health-related activities. Purposive sampling ensured inclusion of information-rich participants able to provide detailed perspectives on unmet needs affecting workers, students, and surrounding communities.Recruitment was structured to achieve relative homogeneity within FGDs to facilitate open discussion and reduce power imbalances, while KIIs targeted individuals with specialized knowledge, leadership roles, or institutional responsibilities relevant to workplace health, school health, service delivery, and community wellbeing.

### Data Collection Methods

A total of three FGDs were conducted. Each FGD comprised participants drawn from similar professional or institutional backgrounds to promote shared understanding and balanced discussion. FGDs explored perceived unmet needs and health challenges, including requests for basic supplies and services related to WASH, first aid preparedness, occupational safety, sexual and reproductive health, preventive screening, and psychosocial wellbeing, as well as broader contextual constraints affecting access to care. FGDs were guided by a semi-structured discussion guide covering domains such as priority health concerns, barriers to care, unmet service needs, gaps in workplace/school/community support, and suggested resources or interventions. Discussions continued until thematic saturation was achieved, defined as the point at which no new substantive themes emerged. Key informant interviews were conducted with individuals identified as having direct experience in organising, delivering, supervising, or responding to health needs in the study setting. These included school administrators, nurses, health and safety officers, supervisors, and other institutional leaders. KIIs enabled deeper exploration of health system gaps, institutional challenges, and service-related unmet needs affecting workers and students. KIIs were conducted until saturation was reached. All FGDs and KIIs were conducted at pre-arranged times in private, quiet locations that ensured confidentiality and minimised interruptions. All sessions were audio-recorded with participants’ consent. Field notes were taken during and immediately after each session to capture contextual observations, group dynamics, and non-verbal cues. Audio recordings were transcribed verbatim, and transcripts were reviewed for completeness and accuracy prior to analysis.

### Data Analysis

Data were analysed using an inductive thematic analysis approach, allowing themes to emerge directly from participants’ narratives rather than being imposed a priori. Analysis involved several iterative stages: familiarisation through repeated reading of transcripts; open coding to identify meaningful units of text; grouping of codes into broader categories; and refinement of categories into overarching themes. Analysis focused on identifying patterns related to unmet basic healthcare needs and broader health challenges within Namanve Industrial Park and surrounding communities, including gaps in preventive services, access to essential commodities, WASH infrastructure, occupational safety systems, sexual and reproductive health support, and mental health and psychosocial services. Representative quotations were selected to illustrate each theme. To enhance trustworthiness, multiple strategies were employed. Triangulation was achieved through use of both FGDs and KIIs and inclusion of participants from diverse stakeholder groups across industrial and school settings. Saturation guided data collection to ensure depth and completeness of themes. An audit trail of transcripts, codes, and thematic decisions was maintained throughout analysis. Reflexive discussions within the research team helped minimise individual bias and strengthen interpretive validity.

## Results: Unmet Health Needs in Namanve Industrial Park and Surrounding Communities

Analysis of focus group discussions and key informant interviews revealed multiple, interrelated unmet health needs affecting workers, students, and surrounding communities in and around Namanve Industrial Park. Participants frequently expressed these unmet needs through requests for materials, services, and infrastructure, which collectively reflect systemic gaps in basic healthcare provision, prevention, and psychosocial support. The findings are organised into six major thematic domains.

### Theme 1: Unmet Needs in Basic Preventive and Primary Healthcare Services

Participants consistently reported limited access to routine screening, testing, and primary healthcare services. Requests included HIV testing and treatment support, cervical and prostate cancer screening, diabetes testing, malaria diagnostics, immunisation services (including HPV vaccination), and general medical check-ups.

Several respondents highlighted that health services are often accessed late, when conditions have already progressed, due to cost, distance, or lack of organised workplace or school-based services. One key informant noted that workers and students frequently present with advanced illness because “there is no routine screening; people only come when they are already very sick.”

Conditions commonly mentioned included malaria, typhoid, peptic ulcer disease (often linked to *Helicobacter pylori*), urinary tract infections, sexually transmitted infections, HIV, diabetes, hypertension, and respiratory tract infections. Participants emphasised that many of these conditions are preventable or manageable if early detection and basic services were accessible.

### Theme 2: Unmet Needs in Sexual, Reproductive, and Maternal Health

Sexual and reproductive health needs were among the most frequently cited unmet needs, particularly for women and adolescent girls. Participants repeatedly requested sanitary pads, family planning services, contraceptives, condoms, antenatal care, and maternal support items, including mama kits.

Several key informants described high rates of unplanned pregnancies, unsafe abortions, and teenage pregnancies, especially among casual workers and students living in congested settlements. One participant explained that “some fear family planning, and they end up pregnant,” while another described girls presenting with bleeding due to complications from unsafe abortions.

Menstrual hygiene was a major concern in both schools and workplaces. Lack of pads, disposal facilities, incinerators, and washing spaces resulted in absenteeism, discomfort, and stigma. Participants stressed that without these basic provisions, girls’ participation in education and work is compromised.

### Theme 3: Unmet Needs in First Aid, Occupational Safety, and Emergency Preparedness

A major theme across industrial and school settings was the absence of functional first aid systems. Participants requested first aid kits, medications (including antiseptics, cotton wool, and spirits), gloves, masks, helmets, boots, protective clothing, and other personal protective equipment (PPE).

Common injuries reported included cuts, burns, chemical splashes (including caustic substances to the eyes), falls, musculoskeletal injuries, and occasional accidents. Participants noted that injuries were often managed inadequately due to lack of supplies or trained personnel.

One informant highlighted that “injuries are common, even if major accidents are not,” emphasising the need for preparedness rather than reactive care. The lack of designated sickbays, examination beds, and mandatory workplace clinics further compounded emergency response challenges.

### Theme 4: Unmet Needs in Water, Sanitation, Hygiene, and Environmental Health

WASH-related unmet needs were repeatedly mentioned across interviews and discussions. Participants requested soap, detergents, handwashing facilities, water dispensers, drinking water bottles, soap dispensers, sanitisers, dustbins, segregation bins, and water filtration equipment.

Environmental health concerns included poor waste management, lack of bins, stagnant water creating mosquito breeding sites, congestion in dormitories and slum settlements, and inadequate bathing facilities. Several participants linked these conditions to malaria, diarrhoeal diseases, skin infections, urinary tract infections, and respiratory illnesses.

Environmental aesthetics and behaviour change were also highlighted, with requests for signage, posters, painted health messages, tree planting, and structured waste disposal systems to promote healthier surroundings.

### Theme 5: Unmet Needs in Mental Health and Psychosocial Support

Mental health emerged as a cross-cutting and deeply entrenched unmet need, particularly among students and young workers. Participants described high levels of stress, anxiety, emotional instability, trauma, substance use, and behavioural disturbances.

Key informants cited academic pressure, family breakdown, poverty, exploitation, social media exposure, cyber risks, substance abuse, bullying, body image concerns, and lack of parental supervision as major contributors. Several alarming incidents were described, including self-harm, hysteria-like episodes (“shake-shake”), emotional breakdowns after relationship conflicts, and suicidal ideation following abortion complications.

Participants repeatedly emphasised the absence of counselling services, social workers, and structured psychosocial support systems, especially in schools. One informant stated that students “need to be heard and encouraged,” while another advocated for permanent or visiting social workers to support vulnerable learners.

### Theme 6: Unmet Needs Linked to Living and Working Conditions

Participants highlighted that many health problems are driven by structural and socioeconomic conditions rather than individual behaviours alone. Casual workers earning low daily wages were described as living in congested slum areas such as Kileku, Nantabulirwa, Kolo, and Buwam, characterised by poor sanitation, overcrowding, and limited access to services.

Health issues linked to these conditions included respiratory infections, gastrointestinal illnesses, malaria, musculoskeletal pain from prolonged standing or heavy labour, poor nutrition, obesity in some groups, and malnutrition in others. Lack of medical insurance for casual workers was repeatedly cited as a major barrier to accessing care.

Participants stressed that without addressing living and working conditions—through better housing, sanitation, occupational protections, and employer engagement—health education alone would be insufficient.

## Discussion

This qualitative study provides an in-depth understanding of unmet basic health needs within an industrial and peri-urban context and demonstrates that health challenges in Namanve Industrial Park and its surrounding communities are not isolated clinical problems, but manifestations of interconnected structural, occupational, and social vulnerabilities. The findings show that participants’ requests for health-related donations—such as sanitary pads, first aid kits, soap, water facilities, screening services, and personal protective equipment—are not expressions of dependency, but indicators of persistent gaps in access to essential preventive and promotive health services. Similar qualitative studies increasingly recognise that material requests in low-resource settings function as diagnostic signals of system failure rather than individual neediness (Kruk et al., 2021; Hanson et al., 2023).

## Structural and Systemic Drivers of Unmet Health Needs

A central insight from this study is that unmet health needs are deeply rooted in systemic limitations of health service delivery within industrial and peri-urban environments. Although government health facilities are geographically present around Namanve, participants described barriers related to affordability, service responsiveness, time compatibility, and relevance to industrial and school contexts. The repeated demand for basic WASH and reproductive health commodities reflects persistent implementation gaps between national policy commitments and on-the-ground service availability. Recent health systems research confirms that proximity to facilities alone does not ensure access, particularly for populations facing informal employment, rigid work schedules, and indirect user costs (Barasa et al., 2021; Roberts et al., 2022). These findings reinforce the need to reconceptualise access as a function of usability and system alignment, rather than physical availability alone.

### Occupational Vulnerability and Informal Employment

The study highlights how informal and casual employment arrangements intensify unmet health needs and undermine preventive care. Workers without insurance, job security, or paid sick leave described delayed care-seeking, self-medication, and reliance on first aid or donations. The strong demand for first aid kits, on-site testing, and protective equipment aligns with global evidence showing that occupational health risks in low- and middle-income countries are often transferred from employers to workers, particularly in loosely regulated industrial settings (International Labour Organization, 2021; Rantanen et al., 2022). Importantly, the findings suggest that unmet needs are not due to lack of awareness of occupational risks, but to institutional arrangements that prioritise productivity over worker wellbeing—a pattern increasingly documented in peri-urban industrial zones across Africa and Asia (Loewenson et al., 2023).

### Gendered Health Needs and Reproductive Vulnerability

The prominence of sanitary pads, family planning services, maternal supplies, and cervical cancer screening reveals pronounced gendered dimensions of unmet health need. Female workers and students face intersecting risks related to menstruation management, unintended pregnancy, unsafe abortion, and limited access to reproductive health services. Recent global reviews demonstrate that inadequate menstrual hygiene management contributes directly to absenteeism, psychosocial distress, and poor educational and workplace outcomes, particularly in low-income and peri-urban settings (Hennegan et al., 2021; Sommer et al., 2022). The study’s findings further highlight how stigma, cost barriers, and service fragmentation continue to undermine uptake of reproductive health services, reinforcing gendered inequities within industrial–community interfaces.

### Mental Health as a Silent but Central Unmet Need

A particularly significant contribution of this study is the identification of mental health as a pervasive yet largely unaddressed unmet need among both workers and students. Participants described stress, anxiety, trauma, low self-esteem, substance use, behavioural crises, and psychosomatic symptoms that were often managed informally or misclassified as disciplinary or medical issues. This aligns with emerging evidence that mental health conditions in industrial and peri-urban populations are frequently under-diagnosed and poorly integrated into primary healthcare and school health systems (Patel et al., 2022; WHO, 2022). The narratives of emotional distress, self-harm risk, and social instability among adolescents underscore the urgency of integrating psychosocial support and counselling services into non-clinical settings such as schools and workplaces.

### Environmental and Living Conditions as Determinants of Health

Participants consistently linked disease patterns—including malaria, typhoid, respiratory infections, skin infections, and gastrointestinal illnesses—to poor environmental conditions in surrounding informal settlements. Requests for mosquito nets, waste bins, drainage, and environmental signage reflect local recognition of environmental determinants of health. These findings reinforce global urban health evidence showing that infectious disease burden in peri-urban industrial zones is closely tied to inadequate sanitation, overcrowding, and waste management rather than individual behaviours alone (Ezeh et al., 2021; Lilford et al., 2023). Addressing unmet health needs therefore requires coordinated environmental and public health action alongside clinical services.

### Health Information Gaps and Fragmented Preventive Services

Although this article focuses on unmet needs rather than knowledge deficits, the findings reveal that unmet needs are compounded by fragmented and under-resourced health education systems. Requests for posters, brochures, signage, demonstration materials, and digital tools suggest that institutions recognise the importance of health education but lack the infrastructure to sustain it. Recent implementation studies caution that health education without enabling resources can inadvertently widen inequities by increasing awareness without improving capacity to act (Glenton et al., 2021; Peters et al., 2023). This study supports the argument that preventive education must be embedded within service-ready systems to translate knowledge into improved outcomes.

### Implications for Health System Design and Intervention Planning

Collectively, the findings indicate that unmet health needs in industrial and peri-urban settings arise from a misalignment between health system design and the lived realities of vulnerable populations. The convergence of occupational risk, informal employment, gender inequity, mental health stressors, and environmental exposure creates a syndemic burden that cannot be addressed through isolated or facility-centred interventions. The expressed needs clearly point toward demand for on-site, integrated, low-cost, and preventive health services that are compatible with work and school routines.

The findings strongly support the implementation of champion-led, digitally supported health education models, as envisioned in the **“Improving the Basic Health in Uganda Using E-Content”** initiative. However, this study also demonstrates that education alone is insufficient. Effective interventions must pair health education with tangible inputs—essential commodities, screening services, counselling, and environmental supports—if unmet health needs are to be meaningfully reduced. This integrated approach aligns with contemporary global health system reform efforts that emphasise people-centred, context-responsive, and prevention-oriented care models (WHO, 2023).

## Conclusion

Unmet health needs in Namanve Industrial Park and surrounding communities are not incidental but structural. Participants’ requests for material and service support reflect health system gaps that undermine preventive and promotive health efforts. Addressing these unmet needs is critical for designing effective, equitable, and sustainable health interventions in industrial and peri-urban settings.

## Data Availability

The qualitative data underlying the findings of this study consist of audio recordings and verbatim transcripts from focus group discussions and key informant interviews. These data contain potentially identifying information and sensitive personal narratives and therefore cannot be shared publicly without restriction. De-identified excerpts supporting the findings are included within the manuscript. Requests for access to anonymized data may be made to the corresponding author, subject to approval by the relevant Research Ethics Committee and the Uganda National Council of Science and Technology (UNCST), and contingent on compliance with ethical and confidentiality requirements.

## Acknowledgements

The authors acknowledge all participants who took part in the focus group discussions and key informant interviews, including industrial workers, supervisors, health and safety personnel, teachers, school administrators, school nurses, and community members from Namanve Industrial Park and surrounding communities. We also acknowledge the leadership of participating industries and schools for granting permission and providing supportive environments for data collection.

## Ethical Considerations

Ethical approval for this study was obtained from the relevant institutional Research Ethics Committee. Permission to conduct the study was also obtained from the Uganda National Council of Science and Technology (UNCST). All study procedures were conducted in accordance with national and institutional ethical guidelines for research involving human participants. No personal identifiers were collected, and all data were anonymised prior to analysis.

## Consent Statement

Written informed consent was obtained from all participants prior to participation in focus group discussions and key informant interviews. Participants were informed about the purpose of the study, the voluntary nature of participation, and their right to withdraw at any time without any consequences. Consent also included permission to audio-record discussions and interviews. No identifying information was collected or reported.

## Patient and Public Involvement

Patients and members of the public were not involved in the design of the study. However, members of the public—including workers, students, and community members—were actively involved as participants in the data collection process through focus group discussions and key informant interviews. Their perspectives directly informed the study findings. The study did not involve patient-level clinical data or patient contact in healthcare settings. Findings from this study will be shared with participating institutions and stakeholders to inform the design and implementation of context-appropriate health education interventions that benefit workers, students, and surrounding communities.

## Funding

The funding was obtained from KOFIH-Korean Foundation For international healthcare.

## Notes

### Competing Interest Statement

The authors have declared no competing interest.

### Funding Statement

The author(s) received no specific funding for this work.

### Author Declarations

This study received ethical approval from the Uganda Christian University REC (UCUREC2025-1979). Administrative clearance to conduct the study was also obtained from the Uganda National Council of Science and Technology (UNCST)HS6818ES. The research was conducted in accordance with national and institutional ethical guidelines and the principles of the Declaration of Helsinki. Written informed consent was obtained from all participants prior to participation in focus group discussions and key informant interviews. Participants were informed about the purpose of the study, the voluntary nature of participation, and their right to withdraw at any time without any consequences. Permission to audio-record discussions and interviews was obtained from all participants. No personal identifiers were collected. All data were anonymised prior to analysis, and confidentiality of participants was strictly maintained throughout data collection, analysis, and reporting.

